# The progressive loss of brain network fingerprints in Amyotrophic Lateral Sclerosis predicts clinical impairment

**DOI:** 10.1101/2022.01.28.22270004

**Authors:** Antonella Romano, Emahnuel Trosi Lopez, Marianna Liparoti, Arianna Polverino, Roberta Minino, Francesca Trojsi, Simona Bonavita, Laura Mandolesi, Carmine Granata, Enrico Amico, Giuseppe Sorrentino, Pierpaolo Sorrentino

## Abstract

Amyotrophic lateral sclerosis (ALS) is a neurodegenerative disease characterised by functional connectivity alterations in both motor and extra-motor brain regions. In network analysis, fingerprint represent a valid approach able to assess the subject-specific connectivity features of a given population. Here, we applied the Clinical Connectome Fingerprint (CCF) analysis to source-reconstructed magnetoencephalography (MEG) signals in a cohort of seventy-eight subjects: thirty-nine ALS patients and thirty-nine healthy controls. We set out to develop an identifiability matrix to assess the extent to which each subject was recognisable based on his/her connectome. The analysis was performed in all five canonical frequency bands. Then we built a multilinear regression model to test the ability of “clinical fingerprint” to predict the clinical evolution of disease assessed by the Amyotrophic Lateral Sclerosis Functional Rating Scale-Revised (ALSFRS-r), the King’s disease staging system and the Milano-Torino Staging (MiToS) disease staging system. We found a drop in patients’ identifiability in the alpha band compared to the healthy controls. Furthermore, the “clinical fingerprint” was predictive of ALSFRS-r (p=0.0397; β=32.8), King’s (p=0.0001; β= -7.40) and MiToS (p=0.0025; β= -4.9) scores and negatively correlated with King’s and MiToS scales according to Spearman’s correlation. Our results demonstrated the ability of the CCF approach in assessing the individual motor condition and its relationship with ALS disease. Thanks to the subject-specific characteristic of this approach, we hope that further exploration related to its clinical application may help improve the management of disease.

## INTRODUCTION

Since Charcot’s first comprehensive anatomo-clinical description in 1874 (Rowland, 2001), Amyotrophic Lateral Sclerosis (ALS) has been defined as a neurodegenerative disease affecting both the upper and lower motor neurons. More recently, clinical, molecular, and neuroimaging evidence suggest that ALS is a pathology that spreads well beyond the motor system (Agosta et al., 2016; Bersano et al., 2020). Besides motor impairment, almost 50% of ALS patients show extra-motor symptoms which involve the cognitive (Goldstein and Abrahams, 2013) and behavioural (Phukan et al., 2007) domains, resulting in a clinical picture that overlaps with frontotemporal dementia (FTD) (Lomen-Hoerth et al., 2002). This overlapping is also confirmed by the presence of TDP-43 protein in several brain regions in both ALS and FTD patients (Geser et al., 2008; Rademakers et al., 2012).

In this regard, neuroimaging techniques, such as f-MRI, PET, and SPECT have been used to investigate functional connectivity modifications in ALS patients (Trojsi et al., 2012; Turner et al., 2012). Mohammedi et al., found that the resting-state functional connectivity alterations mainly occur in the default mode and in the sensori-motor networks (Mohammadi et al., 2009). Interestingly, these changes were correlated with the decline of the motor functions (Fraschini et al., 2018). Similarly, Verstrate et al., through f-MRI analysis, highlighted increased functional connectivity in the motor network which was reflected in a more rapid disease progression (Verstraete et al., 2011). In addition, a magnetoencephalography (MEG) study underlined a widespread topological reorganisation of the brain network in the ALS, which becomes more connected as the disease progresses (Sorrentino et al., 2018). Taken together, these observations highlight the possible role of brain functional connectivity analysis in monitoring disease progression (Iturria-Medina and Evans, 2015), and consequently in refining diagnosis, improving clinical management, and setting up biomarkers for testing progression-modifying drugs. However, the predictive capacity of these approaches in assessing the individual clinical condition is still largely elusive.

Fingerprinting analysis, deriving from functional connectomes (FC), is a methodological approach able to define the subject-specific characteristics of each individual (Amico and Goñi, 2018; Finn et al., 2015; Sareen et al., 2021). It has been tested in both clinical and healthy populations, revealing that individuals affected by neurodegenerative diseases showed a faded identifiability with respect to healthy individuals (Sorrentino et al., 2021b; Svaldi et al., 2021). Interestingly, the reduced identifiability was able to predict the subject-specific clinical features of patients, leading to the concept of *Clinical Connectome Fingerprint* (CCF) (Sorrentino et al., 2021b). In particular, investigating the MEG-based FCs of subjects affected by mild cognitive impairment (MCI) and healthy controls, Sorrentino et al. (Sorrentino et al., 2021b) built an identification score based on the similarity between FCs of patients and healthy subjects (I-clinical), which predicted the patients’ individual cognitive decline.

In the present study, we hypothesised that the global rearrangement of the brain connectivity occurring in ALS (Menke et al., 2017; Sorrentino et al., 2018; Zhou et al., 2016) led to a reduction in brain identifiability that, in turn, may be related to the clinical severity of disease. Hence, we believe that the CCF methodology could be profitably exploited to extract subject-specific connectome features useful in predicting the individual clinical impairments in ALS patients. In order to test our hypothesis, we applied the CCF to source-reconstructed magnetoencephalography (MEG) data in a cohort of seventy-eight subjects, including thirty-nine ALS patients and thirty-nine healthy controls. The functional connectomes were obtained through the phase linearity measurement (PLM), a metric that estimates the synchrony level between pairs of MEG signals(Baselice et al., 2018). Subsequently, we assessed the identifiability rate for both patients and controls within the five canonical frequency bands, expecting that the patients’ FCs would be less recognisable. Finally, to determine whether the identifiability degree of the patients was linked to the clinical condition, we calculated the I-clinical score, which provides individual information about the similarity of each patient’s FC to the average FC of the control group. We tested the predictive ability of the I-clinical with respect to three main clinical ALS scales. In particular we examined Total Amyotrophic Lateral Sclerosis Functional Rating Scale-Revised (ALSFRS-r) which is used to assess the patients’ clinical status (Cedarbaum et al., 1999), and King’s (Balendra et al., 2019) and Milano-Torino Staging (MiToS) (Fang et al., 2017) disease staging systems, which both take into account the chronological onset of symptoms during the disease course.

## METHODS

### Participants

Thirty-nine ALS patients (29 males and 10 females; mean age 59.63; SD ± 12.87; mean education 10.38 years SD±4.3) were recruited for the present study. The ALS diagnosis agreed with the revised El-Escorial criteria(Brooks, 1994) of ALS and none of selected patients showed a mutation in any of the following genes: SOD1, TARDBP, FUS/TLS, and C9ORF72. Inclusion criteria were: 1) no major medical illness and no use of substances that could interfere with MEG signals; (2) no other major systemic, psychiatric, or neurological diseases; and (3) no focal or diffuse brain damage reported at MRI assessment. The control group was composed of thirty-nine subjects as well (28 males and 11 females) matched for age (64±10.4) and education (12±4.3). The patients’ group underwent a neurophysiological and motor screening (see Table 1). Total Amyotrophic Lateral Sclerosis Functional Rating Scale-Revised (ALSFRS-r) was used to assess the preservation of the patient’s physical functions: the greater the score, the better the patient’s condition; King’s disease staging system in which the higher the stage the worse is the patient’s clinical condition and the Milano-Torino staging system (MiToS) which is composed of six stages based on the functional impairment of the patient assessed by ALSFRS-r. The study protocol was approved by the Local Ethics Committee (University of Campania “Luigi Vanvitelli”) with the protocol number 591/2018, and all participants provided written informed consent in accordance with the Declaration of Helsinki.

**Table 1:**
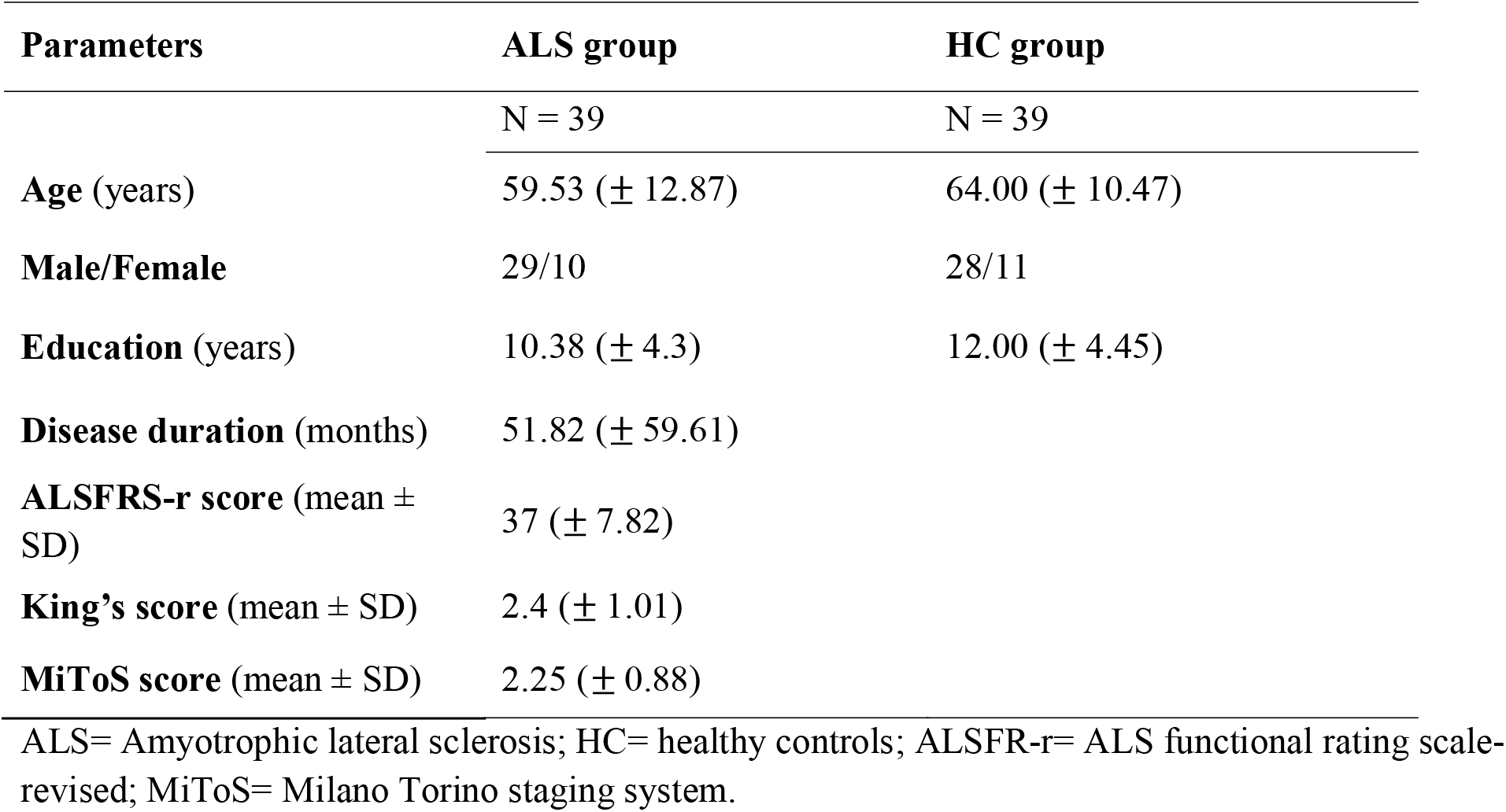
Cohort features. (mean and standard deviation)

### MRI acquisition

MR images of both healthy subjects and patients were acquired on a 3T scanner equipped with an eight-channel parallel head coil (GE Healthcare, Milwaukee, WI, USA). Specifically, three-dimensional T1-weighted images (Gradient-echo sequence Inversion Recovery prepared Fast Spoiled Gradient Recalled-echo, time repetition = 6988 ms, TI = 1100 ms, TE = 3.9 ms, flip angle = 10, voxel size = 1 × 1 × 1.2mm^3^) were acquired. A standard template was used for those participants (six patients and nine healthy controls) who did not complete the MRI in order to obtain the time series of the region of interest (ROIs).

### MEG acquisition

Data were acquired using a MEG system composed of 154 magnetometers SQUID (superconductive quantum interference device) and 9 reference sensors. The acquisition took place in a magnetically shielded room (ATB, Biomag, ULM, Germany) to reduce external noise. To define the position of the head under the helmet we used Fastrack (Polhemus®), that digitised the position of four anatomical landmarks (nasion, right and left pre-auricular points and vertex of the head) and the position of four reference coils (attached to the head of the subject). Each subject was recorded twice (3.5 min each) with a one-minute break, in resting state with closed eyes. Electrocardiography and electro-oculography were also performed to remove physiological artefacts. Finally, after applying an anti-aliasing filter, data were sampled at 1024 Hz.

### Preprocessing

Data preprocessing was performed similarly to Liparoti et al. (Liparoti et al., 2021). In particular, through the implementation of a 4^th^-order Butterworth IIR band-pass filter, using the Fieldtrip toolbox in MATLAB environment, the MEG data were filtered in the band 0.5-48 Hz. Moreover, a Principal component analysis (PCA) was carried out to reduce environmental noise. Finally, we performed an Independent Component Analysis (ICA) in order to remove physiological artefacts.

### Source reconstruction

MEG data were co-registered with the native MRI of each subject. To obtain the time series of the region of interest (ROIs), according to the Automated Anatomical Labeling (AAL) atlas (Gong et al., 2009), we used the volume conduction model proposed by Nolte (Nolte, 2003) and the Linearly Constrained Minimum Variance (LCMV) (Van Veen et al., 1997) beamformer algorithm based on the native MRIs. The time series were canonically filtered in five different frequency band: (i.e., delta (0.5 – 4 Hz), theta (4 – 8 Hz), alpha (8 – 13 Hz), beta (13 – 30 Hz), and gamma (30 – 48 Hz).

### Synchrony estimation

To estimate the synchronisation between brain regions we used the PLM, a measure based on the spectrum of the interferometric signal between any pair of brain regions which is insensitive to volume conduction (Baselice et al., 2018) and ranges from 0 (no synchronisation) to 1 (synchronisation). Hence, using the two separate recordings, we obtained two FCs that we named test and re-test.

### Fingerprint analysis

In order to analyse the individual brain connectivity characteristics of our cohort, we adopted an approach based on the FCs (Figure 1). Firstly, we built an identifiability matrix (IM) for each group, performing a pairwise Pearson’s correlation between test and re-test FCs of our samples. The IM presented the same subjects on both rows and columns, including both test FCs and re-test FCs, respectively (Amico and Goñi, 2018). Each matrix contained information about homo-similarity *(I-self)* represented by the main diagonal, and hetero-similarity (*I-others*) represented by all the elements outside the main diagonal. Then, computing the difference between the mean *I-self* values and the mean *I-other* values we obtained the differential identifiability (*I-diff)*. This score provides an estimation of the fingerprint level of a specific brain data set (Amico and Goñi, 2018).

**Figure 1:**
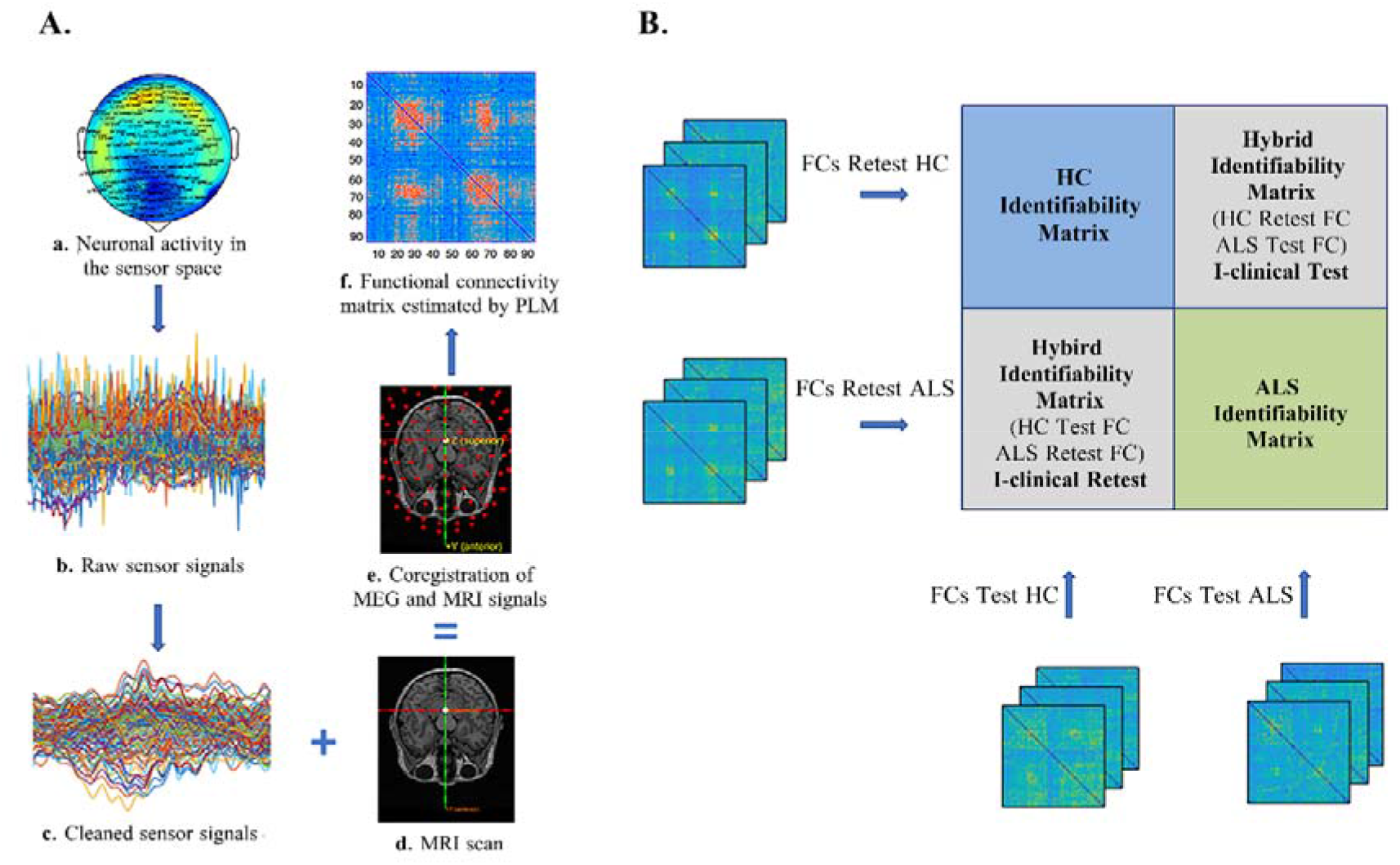
Pipeline analysis and clinical connectome fingerprint application. **(A) a:** the neuronal activity was recorded using a magnetoencephalography (MEG) composed by 154 sensors; **b:** raw MEG signals including noise, and cardiac and blinking artefacts; **c:** MEG signals after removing the noise and the artefacts; **d:** magnetic resonance image (MRI) scan of the subject; **e:** coregistration of MEG and MRI signals to obtain the source reconstruction (beamforming); **f:** functional connectivity matrix estimated by the phase linearity measurement (PLM). **(B)** The blue and the green blocks represents the two identifiability matrices of healthy controls (HC) and Amyotrophic Lateral Sclerosis (ALS) patients, respectively obtained by correlating the test and re-test individual functional connectomes, in each group separately. Crossing the FCs test of the HC with the FCs retest of the ALS and vice-versa we obtained two hybrid identifiability matrices, that allow us to calculate the I-clinical value of each patient (expression of how much an ALS patient is similar to the HC group).

In this study we used an extension of the identifiability approach (Sorrentino et al., 2021b) by crossing the ALS patients and the control group test and re-test FCs to obtain the *I-clinical* score which can assess how much each patient was similar to healthy controls. We built two hybrid matrices: the first one by correlating the controls’ tests with the patients’ re-tests. The second was obtained from a correlation between the controls’ re-tests and the patients’ test. From the average of the values achieved from each hybrid matrix, we found the I-clinical value of each subject (for more detail, see Sorrentino et al.(Sorrentino et al., 2021b).

### Region of interest for fingerprint

To test the edgewise reliability of individual connectomes, we used the intra-class correlation coefficient (ICC) (Koch, 2004), which is a statistical measurement used to assess the separability between elements of the different groups (Amico and Goñi, 2018). In the case of functional connectomes, the edges (i.e the link between two nodes) with a high ICC value are those that contribute the most to identifiability. We evaluated brain fingerprints (Finn et al., 2015) by sequentially adding 50 edges, starting from those that contribute the most to identifiability to those that contribute the least. At each iteration, we calculated the success rate (SR), i.e., the percentage of times that a subject was identifiable with respect to each subject of the same group. We thus obtained a distribution of SR values. To test the reliability of our approach, a null model analysis was performed by adding the edges in random order, one hundred times at each step, to verify that the distribution curve of the SR was statistically meaningful.

### Fingerprint clinical prediction

We tested the hypothesis that the *I-clinical* could predict the clinical impairment of the patient. Thus, we built a multilinear regression model to predict ALSFRS-r, King’s and MiToS scores based on *I-clinical* scores (in the alpha band) alongside with four further predictors: age, sex, education level and duration of disease (expressed in months). Multicollinearity was assessed through variance inflation factor (VIF) (“Identifying influential data and sources of collinearity, ” n.d.) To strengthen the reliability of our approach we performed the leave-one-out-cross validation (LOOCV) (Varoquaux et al., 2017). Specifically, we built *n* multilinear models (where *n* is the number of subjects), excluding each time a different subject from the analysis in order to verify the ability of the model to predict the ALSFRS-r, King’s and MiToS score of the excluded subject.

### Statistical analysis

Statistical analysis was carried out in MATLAB 2021a. I*-self, I-others* and *I-diff* values were compared between the two groups through a permutation test, shuffling the data 10,000 times. At each iteration the absolute difference between the two randomly generated groups was observed, generating a distribution of randomly determined differences. Then, the actual difference between patients and controls was compared to the random distribution to obtain a statistical significance. The possible relationship between *I-clinical* score and clinical condition of the patients was investigated through Spearman’s correlation. Results were corrected by the False Discovery Rate (FDR) method (Benjamini and Hochberg, 1995). Significance level was set at p-value < 0.05 after correction.

## RESULTS

### Fingerprint analysis

As a whole framework, we observed a reduction in ALS patients’ identifiability compared to the HC group in the alpha band (Figure 2). Specifically, we found a statistically significant difference between the *I-self* (pFDR = 0.0249) and the *I-others* (pFDR = 0.0003) of the patients compared to that of the control. We did not find any statistically significant difference between the *I-diff* values between the HC and the ALS groups in any of the five frequency bands.

**Figure 2:**
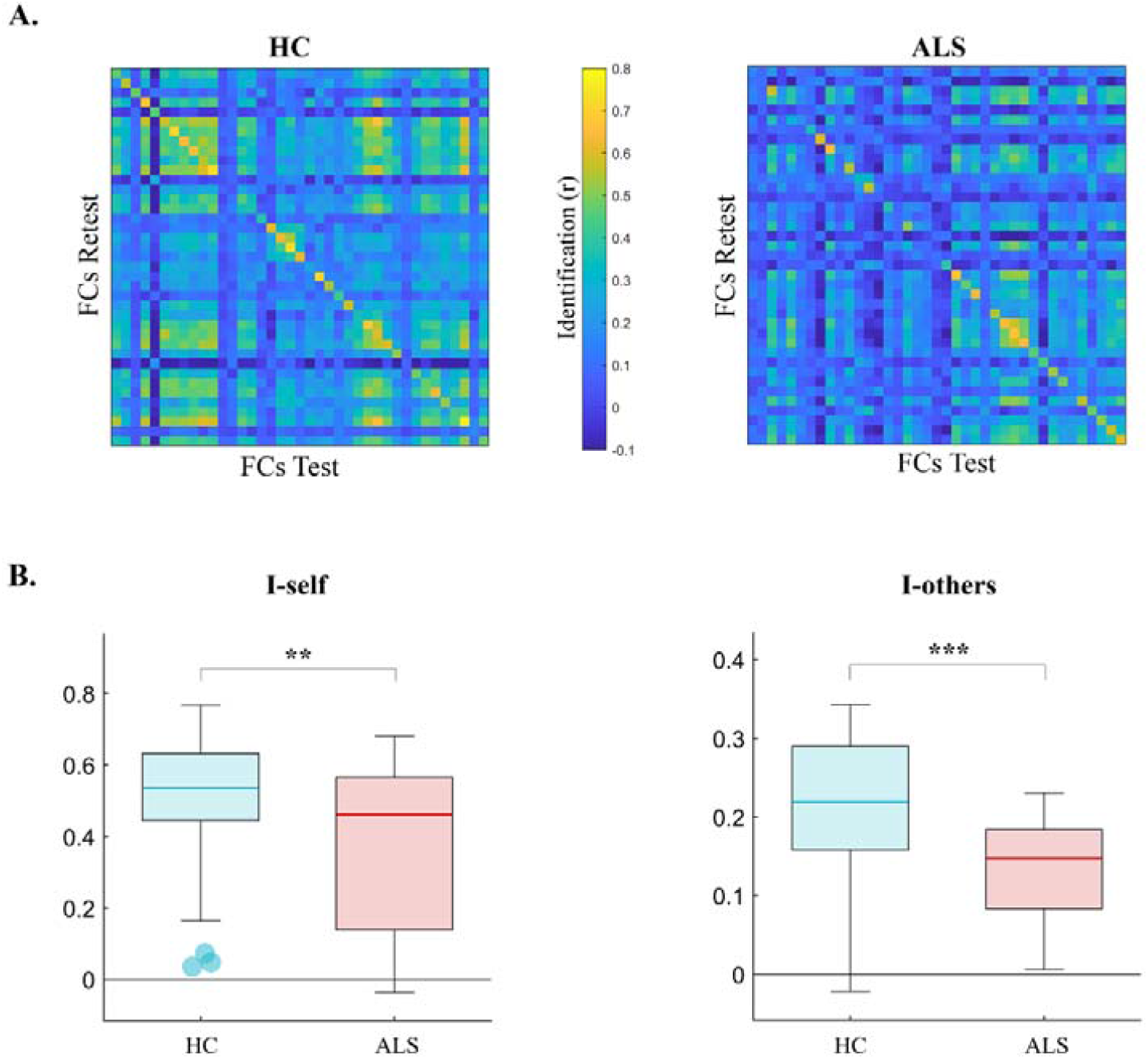
Identifiability matrices and fingerprint features comparison. **(A)**. Identifiability matrices comparison in the alpha band. The main diagonal is representative of the *I-self* while the *I-others* is represented by all the elements outside the main diagonal. The main diagonal of the healthy controls’ (HC) identifiability matrix (IM) is more visible compared to the amyotrophic lateral sclerosis’s (ALS) one, revealing a reduction in homo-similarity (i.e lower I-self values) of the ALS compared to the HC. **(B)** Fingerprint features comparison in the alpha band. HC displays higher values of both I-self and I-others compared to ALS patients. FCs= functional connectomes. significant p value is indicated with **(p <0.01) and *** (p <0.001).

### Edge-based identifiability

In the alpha band we observed that the contribution of brain regions to identifiability was slightly reduced in patients compared to healthy subjects. In fact, in the ICC matrices (Fig. 3A) the control group presented higher values compared to that of the ALS patients revealing a greater stability of the connectivity between the edges. Thus, in the patients the contribution to the fingerprint derived from fewer brain regions compared to controls as shown in Figure 3B.

**Figure 3:**
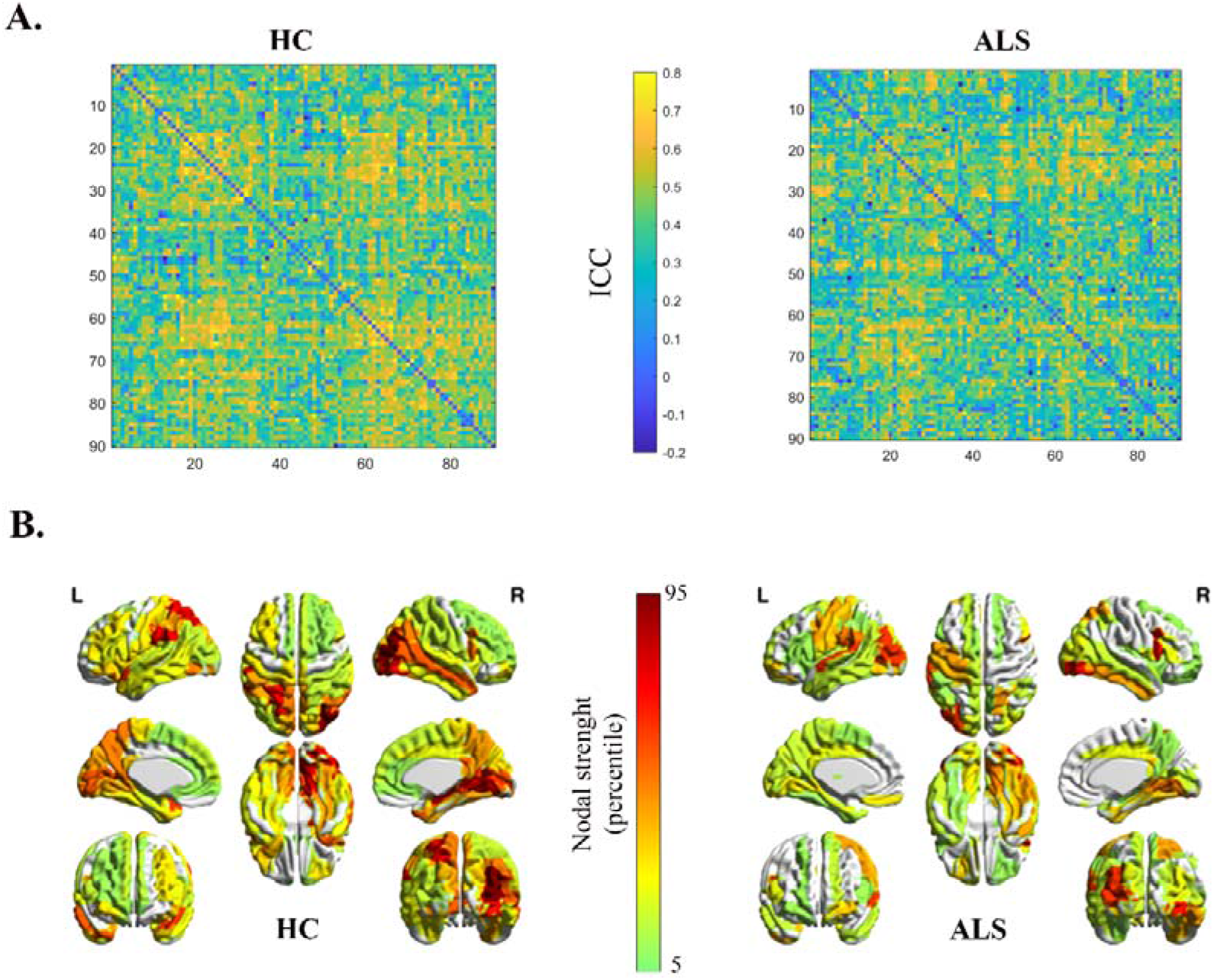
Edge contribution to connectomes’ identifiability. **(A)** The intra-class-correlation (ICC) matrices show the edges contribution to the identifiability in the alpha band. We can observe a drop in ICC determination in the amyotrophic lateral sclerosis (ALS) compared to the healthy controls (HC). This means in the patients there are less reliable brain regions that contribute to self-identification. The **(B)** panel displayed the same results represented as brain renders showing the nodal strength of the most reliable edges. (from 5 to 95 percentile).

Thereafter, we analysed the SR values distribution obtained performing the fingerprint analysis by adding 50 edges at a time, from the most contributing, to the least contributing to fingerprint, according to the ICC matrices. On one hand, the SR curve of the control group immediately reached high values (∼95%) showing a slight, progressively declining value only after exceeding ∼2000 edges. On the other hand, the patients’ SR curve exhibited lower values compared to the healthy group. It reached a plateau between ∼2000 and ∼4500 edges and then progressively declined. Furthermore, the distributions of SR values displayed by the null models of the two groups showed lower values compared to ICC-ordered distributions (Figure 4).

**Figure 4:**
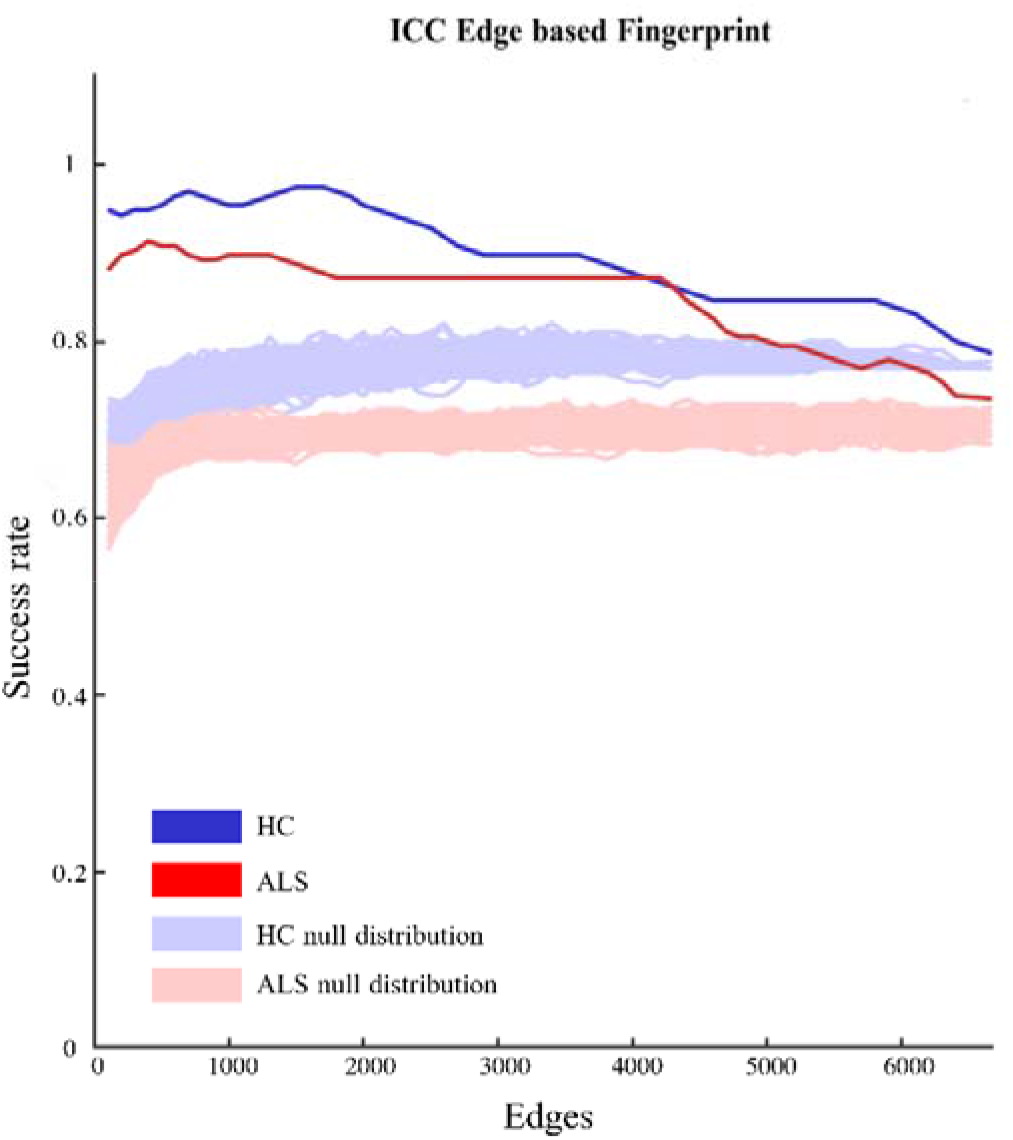
Iterative model of edgewise subjects’ identification. The success rate (SR) distributions of healthy controls (HC, blue line) and Amyotrophic Lateral Sclerosis patients (ALS, red line) were obtained performing the fingerprint analysis by adding 50 edges at a time from those contributing the least to identifiability to those which contributed the most, according to the intraclass correlation (ICC) values. The light blue line and the light red line were representative of the null distribution (obtained by adding the edges in a random order, one hundred times at each step) for the HC and ALS patients respectively. The actual SR distribution displays higher values compared to the null model distributions for both patients and HC.

### Multilinear regression model

In addition, we computed the *I-clinical* score of each ALS patient in the alpha band. We added these values, expression of the similarity of each patient to the healthy group, into a multilinear regression model to test the *I-clinical* capacity to predict the clinical condition assessed by ALSFRS-r, King’s and MiToS clinical scales (Figure 5). Besides the *I-clinical*, the predictive model was composed by four further predictors: age, sex, education level and disease duration. We found that the *I-clinical* significantly predicted the ALSFRS-r (p=0.0397; β=32.8), the King’s (p=0.0001; β= -7.40) and the MiToS (p=0.0025; β= -4.9) scores. The duration of disease significantly contributed to the prediction of ALSFRS-r (p=0.0132; β= -0.0484), King’s (p=0.0015; β=0.0069) and MiToS (p=0.0001; β=0.0077) as well, while no significant contribution of age, education and gender was observed.

**Figure 5:**
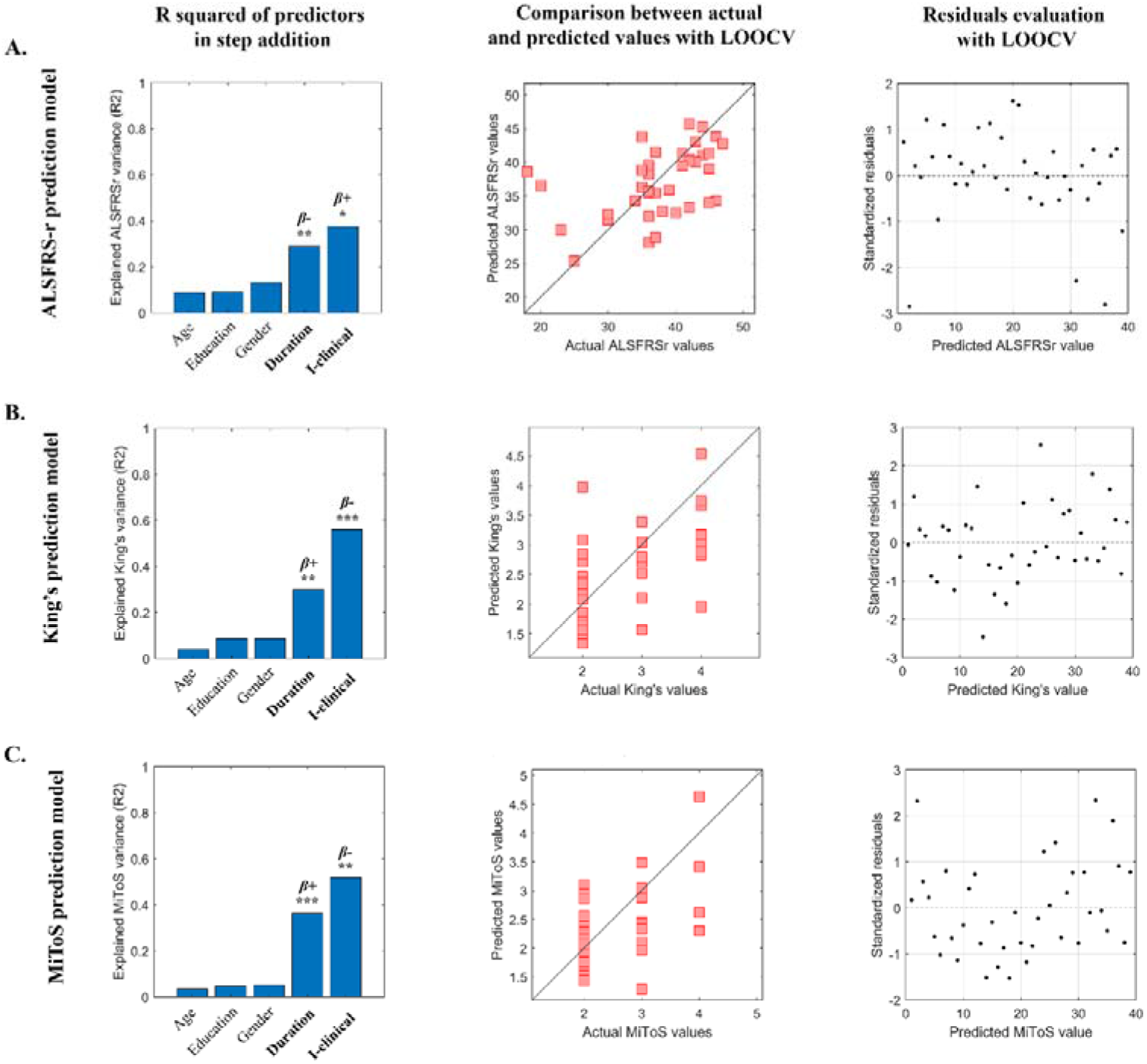
Motor impairment prediction in the alpha band. A multilinear regression model with leave-one-out cross validation (LOOCV) was performed to test the capacity of the “clinical fingerprint” (i.e., the I-clinical score) to predict the motor impairment in amyotrophic lateral sclerosis (ALS) patients. The predictive models of the ALS functional rating scale revised (ALSFRS-r) **(A)**, the King’s disease staging system **(B)** and the Milano Torino staging system (MiTos) are represented on the rows. The left column reports the explained variance obtained by adding the five predictors (age, education, gender, duration of disease and I-clinical in alpha band). The significant predictors are highlighted in bold while the positive and negative coefficients are indicated with β+/β-respectively. The significant p value is indicated with * (p < 0.05) ** (p <0.01) and *** (p <0.001). The central column shows the comparison between the actual and the predicted values of the response variable, validated through LOOCV for **A, B** and **C** respectively. The right column represents the distribution of the standardised residuals (i.e., standardisation of the difference between observed and predicted values) for each row.

### Fingerprint and disease clinical staging

We explored the possibility of a linear relationship between the *I-clinical* and the three above mentioned clinical scores through the Spearman’s correlation in the alpha band. As one can see in Figure 6, we found significant correlations between the I-clinical and both King’s and MiToS disease staging system. Specifically, we found a significant negative correlation, after FDR correction, between *I-clinical* and King’s (r = -0.6041; pFDR = 0.0003) disease staging system and between the *I-clinical* and MiToS (r = -0.4953; pFDR = 0.0040) disease staging system. We did not find any significant correlation between the *I-clinical* and the ALSFRS-r.

**Figure 6:**
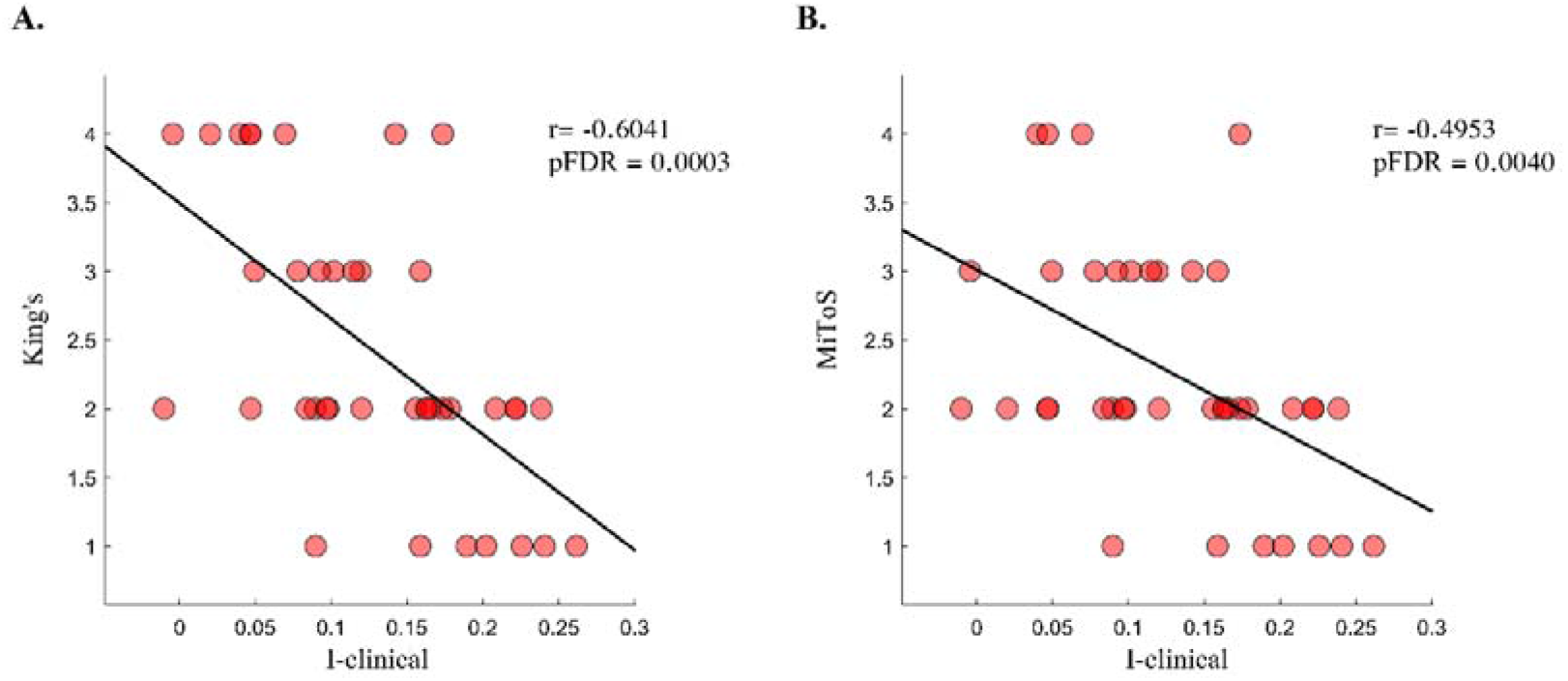
Correlation between motor impairment and I-clinical. **(A)** Spearman’s correlation between the King’s disease staging system and the clinical identifiability (i.e., the I-clinical). The negative correlation coefficient indicates that as the I-clinical scores increase, the King’s scores decrease and thus, the ALS patients show a greater preservation of their motor functions. The **(B)** panel represents the negative correlation between the Milano-Torino staging system (MiTos) and the I-clinical. Higher MiToS scores (worse motor condition) correspond to lower I-clinical scores and vice versa.

## DISCUSSION

In the present study, we applied the clinical connectome fingerprint (CCF) methodology to investigate the subject-specific connectomes features in thirty-nine ALS patients compared to thirty-nine healthy matched controls. Our expectations embraced the idea that the widespread brain connectivity alterations of ALS patients would make the FCs less recognisable with respect to the healthy controls. Furthermore, we tested the hypothesis that such a reduced identifiability might be associated with the individual clinical outcome. Hence, after generating MEG-reconstructed connectomes using the PLM as a synchronisation measure, we proceeded to investigate the identifiability features in ALS patients and healthy individuals. The comparison between the identifiability matrix of the patients and the healthy subjects showed a drop of the identifiability in the patients as compared to the matched controls. Specifically, we found that the patients’ *I-self* and *I-others* were lower than those of the control, while there was no statistically significant difference between the *I-diff* of the two groups. As previously suggested by Sorrentino et al., the loss of identifiability might be ascribed to the large-scale network reorganisation in ALS which make the functional connectomes of the patients less similar to themselves (Sorrentino et al., 2021b). This could be largely due to the alteration of synchrony patterns in the diseased brain (Proudfoot et al., 2019). To this regard, according to Fraschini et al. (Fraschini et al., 2018), ALS patients showed a reduced brain functional connectivity compared to healthy subjects due to the presence of different inter-regional synchrony patterns in the alpha band. Moreover, a MEG study conducted by Proudfoot et al. (Proudfoot et al., 2017), showed that ALS led to a reduction in synchronisation during the execution of motor tasks, arising from the degeneration of neural circuits. Indeed, alterations in synchronisability lead to an impairment of the optimal brain flexibility which in health ensures efficient communication between brain regions (Sorrentino et al., 2021a). Thus, an impairment of brain dynamics implies large-scale communication alterations within the brain, reflecting in a loss of identifiability.

Furthermore, we performed an ICC-based analysis to assess which brain regions mostly contribute to identifiability assuming that the greater the ICC value the greater the contribution of a given edge to identifiability (Amico and Goñi, 2018). As we expected, the reliability of the connectivity between different brain regions was reduced in patients compared to healthy subjects. We hypothesised that this result might be ascribed to the widespread functional alterations which contributed to determine a loss of a stable subject-specific connectivity in ALS patients. Several hypotheses have been put forward concerning the occurrence of these alterations. According to Poujois et al., this process might be caused by a reorganisation of both the motor and extra-motor regions through neural plasticity modulations (Poujois et al., 2013) which occurs as a compensatory mechanism of ALS neurodegeneration (Trojsi et al., 2012). In another study proposed by Turned et al., it has been shown that the ALS brain functional alterations could result from the loss of inhibitory interneurons (Turner and Kiernan, 2012). In this regard, note that a reduction in GABAergic inhibition is responsible for cortical hyperexcitability typical of ALS (Kiernan and Petri, 2012) and, more importantly, the reduced cortical inhibition, ascribed to the loss of GABAergic interneurons, seemed to be responsible for an increase in functional connectivity (Douaud et al., 2011).

Following this, we investigated the contribution of the edges in identifying the FCs of both groups. As shown in Figure 4, a few hundred edges were sufficient to obtain optimal subject recognition. On the contrary, increasing the number of edges did not improve identification, rather it was reduced. We hypothesised that this finding is a consequence of the fact that subject-specific patterns are encoded within a restricted number of connections. Conversely, including the whole connectome we are more likely to include patterns that are common among different individuals (Gratton et al., 2018). Furthermore, the downward trend in edge-based identifiability was even more prominent in patients than in controls. This is in accordance with our results on the reliability of the connections, which in patients displayed a reduced number of stable links between brain areas.

Finally, we explored the relationship between the CCF features and the clinical assessment of the disease and its rate progression. Previous evidence suggested that alterations in functional connectivity can be considered as a marker of disease progression (Dubbelink et al., 2013) and they can be related to the rate of disability (Fraschini et al., 2016). Our results are in agreement with the above findings and define a new perspective that may be helpful in the management of the disease. Indeed, our analysis highlighted an association between the subject-specific clinical identification (i.e., *I-clinical*) and the motor impairment evaluation. Specifically, the multilinear regression model that we built pointed out that, among all the predictors (age, sex, education level and duration of disease), the *I-clinical*, together with the duration of the disease, was able to significantly predict the ALSFRS-r, King’s disease staging system and MiToS scores. Furthermore, the generalisation and prediction capacity of our models was bolstered by LOOCV. With regard to the disease duration, it is obvious that such a relation would exist, since ALS is a neurodegenerative disease that worsens over time. On the contrary, the result represented by the *I-clinical* score prediction brings with it novel information which connect the severity of the disease with the clinical aspects of the brain identifiability. This relationship was strengthened by the negative correlation between the I-clinical and King’s and MiToS scores. Indeed, patients with higher I-clinical scores were more similar to the control group and therefore they showed greater preservation of their motor functions.

## CONCLUSIONS

In summary, in the present paper, we applied the CCF analysis to ALS, to test its reliability in a clinical setting which is predominantly characterised by motor symptoms combined with the presence of cognitive and behavioural symptomatology. As we hypothesised, ALS patients showed reduced identifiability compared to healthy subjects. Moreover, connectomes of patients with higher preservation of their motor functions displayed greater similarity to the connectomes of the healthy individuals. Moreover, the ability of clinical fingerprint to predict the individual rate of disease progression may represent a promising finding in ALS clinical management. Thanks to the subject-specific characteristic of this approach, we hope that further exploration related to its clinical application may help in diagnostic and therapeutic strategies.

## Data Availability

all the data produced in the present study are available upon reasonable request to the authors

## Abbreviations

(ALS): Amyotrophic Lateral Sclerosis
(MEG): Magnetoencephalography
(FC): Functional Connectome
(CCF): Clinical Connectome Fingerprint
(PLM): Phase linearity measurement
(ALSFRS-r): Amyotrophic Lateral Sclerosis Functional Rating Scale-Revised
(MiToS): Milano-Torino Staging disease staging system
(ROIs): Region of interest
(IM): Identifiability matrix
(ICC): Intra-class correlation coefficient
(SR): Success rate
(LOOCV): Leave-one-out-cross validation

## ACKNOWLEDGMENTS

We would like to thank Silvia Lombardini for the english revision of the manuscript.

## DATA AND CODE AVAILABILITY STATEMENT

The data that support the findings of this study are available from the corresponding author upon reasonable request. The data are not publicly available due to the clinical nature of the cohort under study. The code (in MATLAB) used for this analysis is available at the following link: https://github.com/eamico/Clinical_fingerprinting

## AUTHORS CONTRIBUTION

A.R and E.T.L collected and acquired the dataset, processed the data and conceptualised the study; F.T collected the sample; M.L, A.P, R.M, S.B, L.M, C.G, E.A, contributed to interpreting the results and critically revised the article; P.S conceptualised and supervised the study and G.S supervised the study. All authors interpreted the results and wrote the manuscript.

